# Efficacy of Tirzepatide Dual GIP/GLP-1 Receptor Agonist In Patients with Idiopathic Intracranial Hypertension. A Real-World Propensity Score-Matched Study

**DOI:** 10.1101/2024.11.12.24317193

**Authors:** Ahmed Y. Azzam, Muhammed Amir Essibayi, Nathan Farkas, Mohammed A. Azab, Mahmoud M. Morsy, Osman Elamin, Adam Elswedy, Ahmed Saad Al Zomia, Hammam A. Alotaibi, Ahmed Alamoud, Oday Atallah, Hana J. Abukhadijah, Adam A. Dmytriw, Amanda Baker, Deepak Khatri, Neil Haranhalli, David J. Altschul

**Author notes:** **Corresponding Author:** Hana J. Abukhadijah, Medical Research Center, Hamad Medical Corporation, Doha Qatar, P.O. Box 3050.

## Abstract

**Introduction:** Idiopathic intracranial hypertension (IIH) is a neurological disorder characterized by elevated intracranial pressure, predominantly affecting obese women of reproductive age. While GLP-1 receptor agonists have shown promise in IIH management, the potential of dual GIP/GLP-1 receptor activation through tirzepatide remains unexplored. This study aimed to evaluate tirzepatide’s efficacy as an adjunctive therapy in IIH management.

**Methods:** We conducted a retrospective cohort analysis using the TriNetX Global Health Research Network, analyzing data through November 2024. Through propensity score matching, we compared 193 tirzepatide-exposed IIH patients with 193 controls receiving standard care. Primary outcomes included papilledema severity, visual function, headache frequency, and treatment resistance, monitored at multiple follow-up timepoints.

**Results:** Our analysis revealed significant improvements across all measured outcomes in the tirzepatide group. At 24 months, we observed a 68% reduction in papilledema risk (RR 0.320, 95% CI 0.189-0.542, p<0.001), a 73.9% reduction in visual disturbance and blindness risk (RR 0.261, 95% CI 0.143-0.477, p<0.001), and a 19.7% reduction in headache risk (RR 0.803, 95% CI 0.668-0.966, p=0.019). The tirzepatide group demonstrated significant body-mass index reductions, reaching -1.147 kg/m^2^ (95% CI [-1.415, -0.879], p<0.001) at 24 months compared to controls.

**Conclusions:** Our results demonstrate that tirzepatide, when used as an adjunctive therapy, provides significant therapeutic benefits in IIH management, particularly in improving papilledema and visual outcomes. Our findings suggest that dual GIP/GLP-1 receptor activation may offer advantages over traditional single-receptor therapies, potentially through enhanced metabolic regulation and direct effects on intracranial pressure dynamics.

## 1. Introduction

Idiopathic intracranial hypertension (IIH) is a neurological disorder characterized by elevated intracranial pressure (ICP) without an identifiable underlying cause [1, 2]. The condition predominantly affects women of reproductive age with obesity, manifesting through debilitating symptoms including chronic headaches and papilledema, which can lead to permanent visual impairment if left untreated [1, 3]. The pathophysiology of IIH is complex and multifactorial, involving obesity-related mechanisms such as increased intra-abdominal pressure affecting cerebral venous drainage and adipose tissue functioning as an endocrine organ, releasing factors that influence ICP and neuroinflammation [3-5]. Current standard management approaches include medical therapies such as acetazolamide, which reduces cerebrospinal fluid (CSF) production, topiramate, which combines ICP-lowering and migraine prophylaxis effects, and other diuretics [2, 3]. In cases refractory to medical management, surgical interventions including venous sinus stenting, optic nerve sheath fenestration (ONSF), and CSF shunting procedures may be necessary [3, 4, 6].

However, current management options face significant limitations [5, 7]. Medical therapies often produce suboptimal responses, with many patients experiencing persistent symptoms or intolerable side effects that lead to poor compliance [2, 8]. Surgical interventions, while effective in selected cases, carry inherent risks and may require revision procedures [3, 4]. Furthermore, while weight loss has demonstrated effectiveness in IIH management, achieving and maintaining significant weight reduction through lifestyle modifications alone proves challenging for many patients [7], highlighting the need for more effective therapeutic approaches.

Glucagon-like peptide-1 receptor agonists (GLP-1 RAs) have emerged as a promising therapeutic option for IIH [1, 5]. Originally developed for type 2 diabetes management, these agents work by enhancing insulin release, suppressing glucagon, and slowing gastric emptying [1, 5]. Recent evidence has revealed that GLP-1 receptors are present in the brain, including the hypothalamus and choroid plexus, suggesting potential direct effects on CSF production and ICP regulation [1-3]. Clinical studies have demonstrated encouraging results regarding the safety and efficacy of GLP-1 RAs in IIH management [2, 8]. For instance, clinical trials with exenatide showed significant ICP reductions of 5.7 cm CSF within hours of administration, with sustained effects at 12 weeks [2]. Additionally, studies have reported substantial improvements in headache frequency and severity, with 76.9% of patients achieving at least a 50% reduction in monthly headache days when treated with GLP-1 RAs like semaglutide or liraglutide [5, 8].

While current evidence supporting GLP-1 RAs in IIH treatment is promising, studies have primarily focused on exenatide, a short-acting GLP-1 RA [2, 5]. Notably absent from the literature is investigation into tirzepatide’s efficacy in IIH management. Tirzepatide represents a unique dual GIP/GLP-1 receptor agonist with potentially superior weight loss effects and metabolic benefits compared to single-receptor GLP-1 RAs [5]. Given its dual receptor activity and enhanced weight reduction potential, tirzepatide may offer additional benefits for IIH patients through both direct ICP-lowering effects and sustained weight management [4, 5, 8].

This international multicenter study aims to evaluate the efficacy of tirzepatide as an adjunctive therapy to standard IIH management using the TriNetX database [1, 2]. Our retrospective cohort analysis represents the first large-scale investigation of tirzepatide’s role in IIH treatment. The study’s strengths include its substantial sample size, diverse international patient population, and real-world clinical setting, which enhance the generalizability of our findings. By analyzing tirzepatide’s effectiveness as an adjunctive therapy rather than monotherapy, our study design reflects practical clinical scenarios and provides valuable insights into optimizing IIH management strategies [5, 8].

## 2. Methods

### 2.1. Patient Selection and Cohort Establishment

We utilized the TriNetX Global Health Research Network (TriNetX, Cambridge, MA, USA), a federated real-time platform integrating electronic health records from approximately 160 healthcare organizations worldwide [9]. TriNetX network (https://trinetx.com/solutions/live-platform/) encompasses around 197 million patient records across multiple countries, including the United States as the predominant source, along with healthcare data from Australia, Belgium, Brazil, Bulgaria, Estonia, France, Georgia, Germany, Ghana, Israel, Italy, Japan, Lithuania, Malaysia, Poland, Singapore, Spain, Taiwan, United Arab Emirates, and the United Kingdom. Our study leveraged data through November 2024, representing the complete available timeframe in the dataset. The TriNetX platform provides comprehensive patient-level information including demographics, diagnoses, treatments, procedures, and clinical outcomes, coded using standardized medical classification systems such as the International Classification of Diseases, Tenth Revision (ICD-10) and Current Procedural Terminology (CPT) codes. All data within the network are automatically standardized and harmonized to enable consistent analysis across different healthcare organizations and geographical regions. The platform ensures HIPAA compliance through automatic de-identification of patient data while maintaining the integrity of longitudinal records for research purposes.

We utilized the electronic health records to identify adult patients with IIH through the ICD-10-CM classification G93.2. Eligibility requirements included: age 18 years or older at initial diagnosis, verification of IIH diagnosis through documented clinical evaluations, participation in conventional therapeutic protocols, and complete baseline clinical documentation including body mass index (BMI) measurements.

The tirzepatide intervention group was established through pharmaceutical dispensing records (identified via specific RxNorm unique identifiers) indicating tirzepatide administration alongside standard care protocols. The comparison group consisted of patients managed with conventional therapeutics, including but not limited to acetazolamide, topiramate, and weight management strategies, without exposure to any GLP-1/GIP receptor agonists.

Exclusion parameters were comprehensively defined to encompass: secondary intracranial hypertension etiologies (validated through ICD-10 coding), including cerebral venous thrombosis and structural neurological abnormalities; prior utilization of incretin-based therapies within the preceding 12 months; pregnancy or recent post-partum status; insufficient baseline clinical data; and follow-up duration below six months. Incomplete cases were systematically removed from the analysis to ensure data quality.

### 2.2. Propensity Score-Matching

To mitigate selection bias and establish comparable cohorts, we implemented a 1:1 propensity score matching protocol utilizing TriNetX’s built-in analytics features. The propensity score model incorporated multiple variables including demographic characteristics such as age at diagnosis, sex, race (categorized as White, Black or African American, Asian, and Other), and ethnicity (Hispanic or Latino, Non-Hispanic or Latino). Clinical parameters included baseline BMI, comorbidity profiles including endocrine, musculoskeletal, and eye disorders, pre-existing medication usage, and healthcare utilization patterns in the year preceding cohort entry.

The matching procedure employed an optimal matching algorithm with a stringent caliper width of 0.1 standard deviations of the propensity score logit. This refined methodology transformed our initial sample of 438 tirzepatide-exposed patients and 21,654 controls into balanced cohorts of 435 and 870 patients respectively, achieving optimal covariate balance with standardized differences below 0.1 for all matched variables.

### 2.3. Outcome Definition and Monitoring

Primary endpoints were monitored through structured database queries at 3-month, 6-month, 12-month, and 24-month intervals post-treatment initiation. Disease-specific markers included papilledema severity (ICD-10: H47.1), headache frequency and intensity (ICD-10: G44.-), and visual function parameters (ICD-10: H53.-, H54.-). Treatment resistance was defined through a composite of indicators including requirement for escalation of medical therapy, progression to surgical intervention, need for CSF diversion procedures, or optic nerve decompression/ONSF necessity.

Secondary endpoints focused on metabolic parameters, particularly BMI trajectories and body composition changes, documented through standardized clinical measurements at each follow-up interval. These measurements were extracted directly from structured clinical data fields, with both absolute changes and percentage reductions calculated to provide comprehensive assessment of weight-related outcomes.

### 2.4. Statistical Methodology

All analyses were performed using TriNetX’s analytical tools, which employ standardized statistical methodologies. For primary outcomes, we calculated risk ratio/relative risk (RR) with corresponding 95% confidence intervals, and absolute risk differences between groups. Continuous variables were expressed as means ± standard deviations, with between-group comparisons conducted using Student’s t-tests. Categorical variables were analyzed using chi-square tests or Fisher’s exact tests where appropriate. For BMI analysis, we computed both absolute and baseline-adjusted differences between groups, with temporal trends assessed through longitudinal analysis features. Statistical significance was consistently defined as p<0.05, with exact p-values reported where available. Multiple comparison adjustments were applied using the Benjamini-Hochberg procedure to control the false discovery rate.

## 3. Results

We evaluated baseline characteristics between tirzepatide-exposed and control cohorts through propensity score-matching (**Table 1**). Our initial sample comprised 193 patients in the tirzepatide group and 20,683 in the control group, which was subsequently matched to achieve balanced cohorts of 193 patients each. The demographic composition revealed a predominantly female population in both matched groups (91.71% vs 92.75%, p=0.7038). Mean age was comparable between matched cohorts (37.7 ± 8.52 vs 37.5 ± 8.78 years, p=0.8140), though we observed a persistent significant difference in body mass index (BMI) (41.3 ± 8.82 vs 38.7 ± 9.33 kg/m^2^, p=0.0089).

**Table 1.** Baseline Characteristics of Tirzepatide Group and Control Group Before and After Propensity Score Matching for Cohorts.

| Variable | Tirzepatide Before Matching | Control Before Matching | P-value Before | Tirzepatide After Matching | Control After Matching | P-value After |
| --- | --- | --- | --- | --- | --- | --- |
| Sample size, n | 193 | 20,683 | N/A | 193 | 193 | N/A |
| Age (years), mean ± SD | 37.7 ± 8.52 | 35.7 ± 10 | 0.0046 | 37.7 ± 8.52 | 37.5 ± 8.78 | 0.8140 |
| BMI, mean ± SD | 41.3 ± 8.82 | 35.5 ± 9.02 | < 0.0001 | 41.3 ± 8.82 | 38.7 ± 9.33 | 0.0089 |
| Female, n (%) | 177 (91.71%) | 17,633 (85.76%) | 0.0183 | 177 (91.71%) | 179 (92.75%) | 0.7038 |
| Male, n (%) | 10 (5.18%) | 2,504 (12.18%) | 0.0030 | 10 (5.18%) | 10 (5.18%) | 1.0000 |
| **Race/Ethnicity:** |  |  |  |  |  |  |
| White, n (%) | 139 (72.02%) | 12,235 (59.50%) | 0.0004 | 139 (72.02%) | 146 (75.65%) | 0.4176 |
| Black or African American, n (%) | 31 (16.06%) | 3,913 (19.03%) | 0.2955 | 31 (16.06%) | 28 (14.51%) | 0.6713 |
| Hispanic or Latino, n (%) | 11 (5.70%) | 1,924 (9.36%) | 0.0820 | 11 (5.70%) | 14 (7.25%) | 0.5350 |
| Asian, n (%) | 0 (0%) | 299 (1.45%) | 0.0915 | 0 (0%) | 0 (0%) | - |
| **Comorbidities:** |  |  |  |  |  |  |
| Endocrine disorders, n (%) | 186 (96.37%) | 8,554 (41.60%) | < 0.0001 | 186 (96.37%) | 187 (96.89%) | 0.7778 |
| Musculoskeletal disorders, n (%) | 139 (72.02%) | 6,653 (32.36%) | < 0.0001 | 139 (72.02%) | 136 (70.47%) | 0.7358 |
| Eye disorders, n (%) | 130 (67.36%) | 11,466 (55.76%) | 0.0012 | 130 (67.36%) | 126 (65.29%) | 0.6666 |
| Digestive system disorders, n (%) | 122 (63.21%) | 4,897 (23.82%) | < 0.0001 | 122 (63.21%) | 125 (64.77%) | 0.7504 |
| Circulatory system disorders, n (%) | 52 (26.94%) | 3,470 (16.88%) | 0.0002 | 52 (26.94%) | 50 (25.91%) | 0.8174 |

Our matched cohorts demonstrated significant balance across racial and ethnic distributions, with White patients constituting the majority (72.02% vs 75.65%, p=0.4176), followed by Black or African American (16.06% vs 14.51%, p=0.6713) and Hispanic or Latino patients (5.70% vs 7.25%, p=0.5350). Notably, we achieved a balance in comorbidity profiles post-matching, with comparable rates of endocrine (96.37% vs 96.89%, p=0.7778), musculoskeletal (72.02% vs 70.47%, p=0.7358), and eye disorders (67.36% vs 65.29%, p=0.6666). Additional comorbidities, including digestive system disorders (63.21% vs 64.77%, p=0.7504) and circulatory system disorders (26.94% vs 25.91%, p=0.8174), were also well-balanced between groups (**Figure 1**).

**Figure 1.**
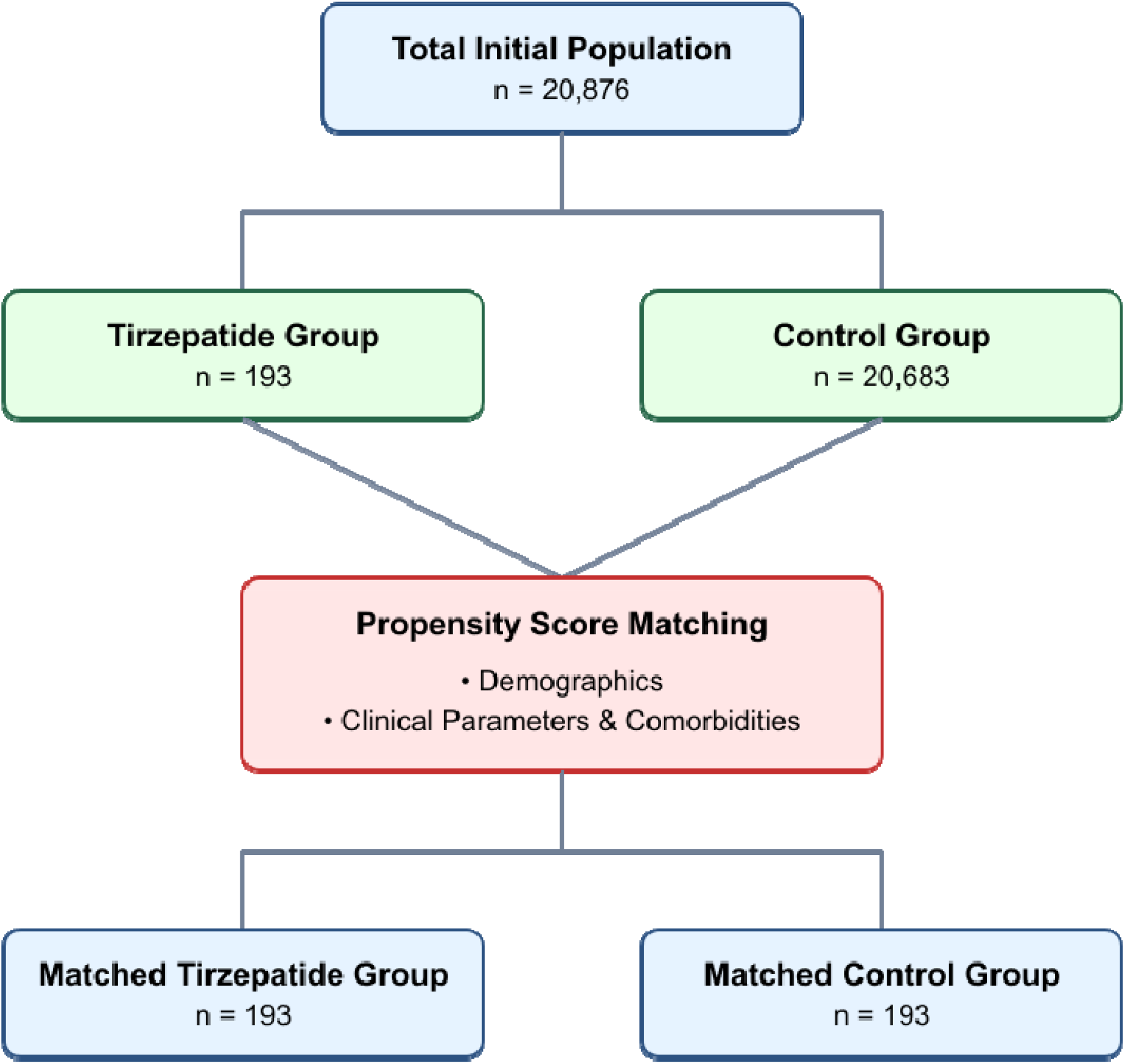
Patients Enrollment Flow Diagram.

### 3.1. Symptoms and Outcomes Analysis

Our analysis revealed significant therapeutic benefits of tirzepatide as an adjunctive therapy across all measured outcomes in patients with IIH (**Table 2**). The most significant improvements were observed in papilledema outcomes, with the tirzepatide group demonstrating a 67.7% reduction in risk at three months (RR 0.323, 95% CI 0.163-0.639, p=0.001). This promising protective effect was maintained throughout the follow-up period, showing a 68% risk reduction at 24 months (RR 0.320, 95% CI 0.189-0.542, p<0.001).

**Table 2.** Outcomes Risk For Tirzepatide Group Compared to Control Group.

| **Outcome** | **Follow-up Timepoint** | **Risk Ratio** | **95% Confidence Interval** | **P-value** |
| --- | --- | --- | --- | --- |
| Papilledema | 3-months | 0.323 | (0.163, 0.639) | 0.001 |
|  | 6-months | 0.342 | (0.188, 0.622) | 0.000 |
|  | 12-months | 0.348 | (0.204, 0.593) | 0.000 |
|  | 24-months | 0.320 | (0.189, 0.542) | 0.000 |
| Headache | 3-months | 0.767 | (0.597, 0.987) | 0.037 |
|  | 6-months | 0.820 | (0.663, 1.015) | 0.066 |
|  | 12-months | 0.849 | (0.697, 1.035) | 0.103 |
|  | 24-months | 0.803 | (0.668, 0.966) | 0.019 |
| Refractory IIH | 3-months | 0.768 | (0.592, 0.997) | 0.046 |
|  | 6-months | 0.813 | (0.650, 1.015) | 0.066 |
|  | 12-months | 0.843 | (0.686, 1.036) | 0.103 |
|  | 24-months | 0.795 | (0.654, 0.965) | 0.019 |
| Visual Disturbances or Blindness | 3-months | 0.385 | (0.191, 0.776) | 0.005 |
|  | 6-months | 0.323 | (0.163, 0.639) | 0.001 |
|  | 12-months | 0.289 | (0.153, 0.549) | 0.000 |
|  | 24-months | 0.261 | (0.143, 0.477) | 0.000 |

Visual outcomes showed similarly significant improvements, with tirzepatide treatment associated with a 61.5% reduction in risk of visual disturbances or blindness at three months (RR 0.385, 95% CI 0.191-0.776, p=0.005). The protective effect strengthened over time, reaching a 73.9% risk reduction at 24 months (RR 0.261, 95% CI 0.143-0.477, p<0.001), representing the most progressive improvement among all measured outcomes.

Headache symptoms showed also statistically significant improvement with tirzepatide therapy. The intervention group demonstrated a 23.3% reduction in headache risk at three months (RR 0.767, 95% CI 0.597-0.987, p=0.037), with sustained benefit showing a 19.7% risk reduction at 24 months (RR 0.803, 95% CI 0.668-0.966, p=0.019).

In cases of refractory IIH, traditionally challenging to manage, the tirzepatide group showed a 23.2% reduction in risk at three months (RR 0.768, 95% CI 0.592-0.997, p=0.046), maintaining a 20.5% risk reduction at 24 months (RR 0.795, 95% CI 0.654-0.965, p=0.019).

The risk difference analysis revealed distinctive longitudinal patterns in treatment response across all outcomes over the different follow-up timepoints in our study (**Figure 2**). The most pronounced therapeutic divergence between tirzepatide and control groups occurred at six months for most outcomes, with sustained benefits through 24 months. This pattern was particularly evident in visual disturbances and papilledema, where risk differences remained consistently significant (p<0.001) throughout the follow-up period. The absolute risk reduction demonstrated clinical significance even at 24 months, with the tirzepatide group maintaining consistently lower risk profiles across all outcomes, as illustrated by the negative risk differences favoring tirzepatide treatment (**Figure 2**).

**Figure 2.**
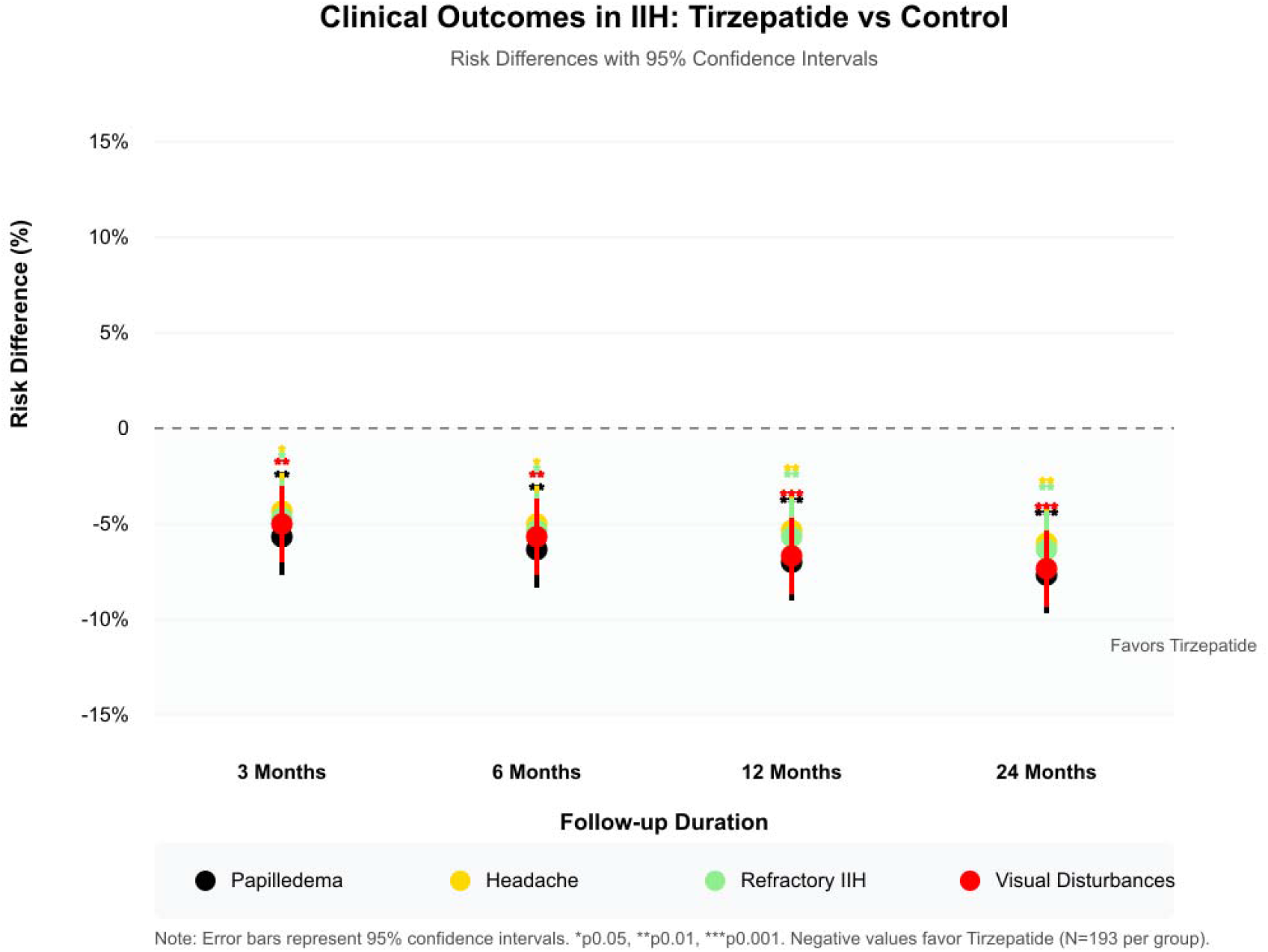
Risk Difference Longitudinal Analysis Over Timepoints For Outcomes.

### 3.2. BMI Longitudinal Analysis

The longitudinal analysis of BMI trends revealed significant differences between the tirzepatide and control groups over the 24-month follow-up period (**Table 3**). At the baseline-adjusted analysis, we observed a progressive BMI reduction in the tirzepatide group, starting with a modest decrease at three months (-0.294 kg/m^2^, 95% CI [-0.581, -0.007], p=0.045). This reduction became more pronounced at subsequent timepoints, reaching -0.733 kg/m^2^ at six months (95% CI [-1.012, -0.454], p<0.001), -0.986 kg/m^2^ at 12 months (95% CI [-1.259, -0.713], p<0.001), and achieving maximal effect at 24 months with -1.147 kg/m^2^ (95% CI [-1.415, -0.879], p<0.001).

**Table 3.**
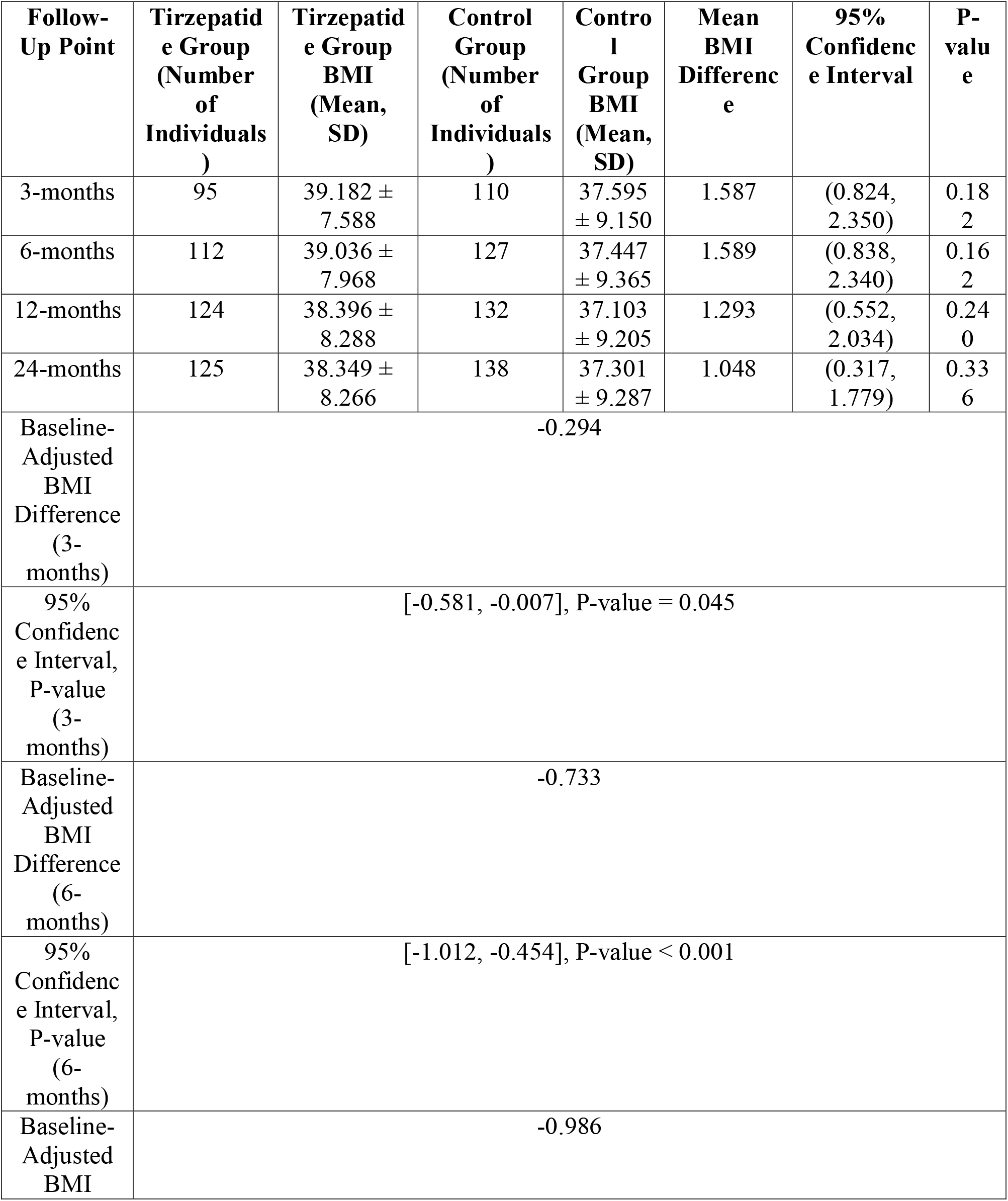
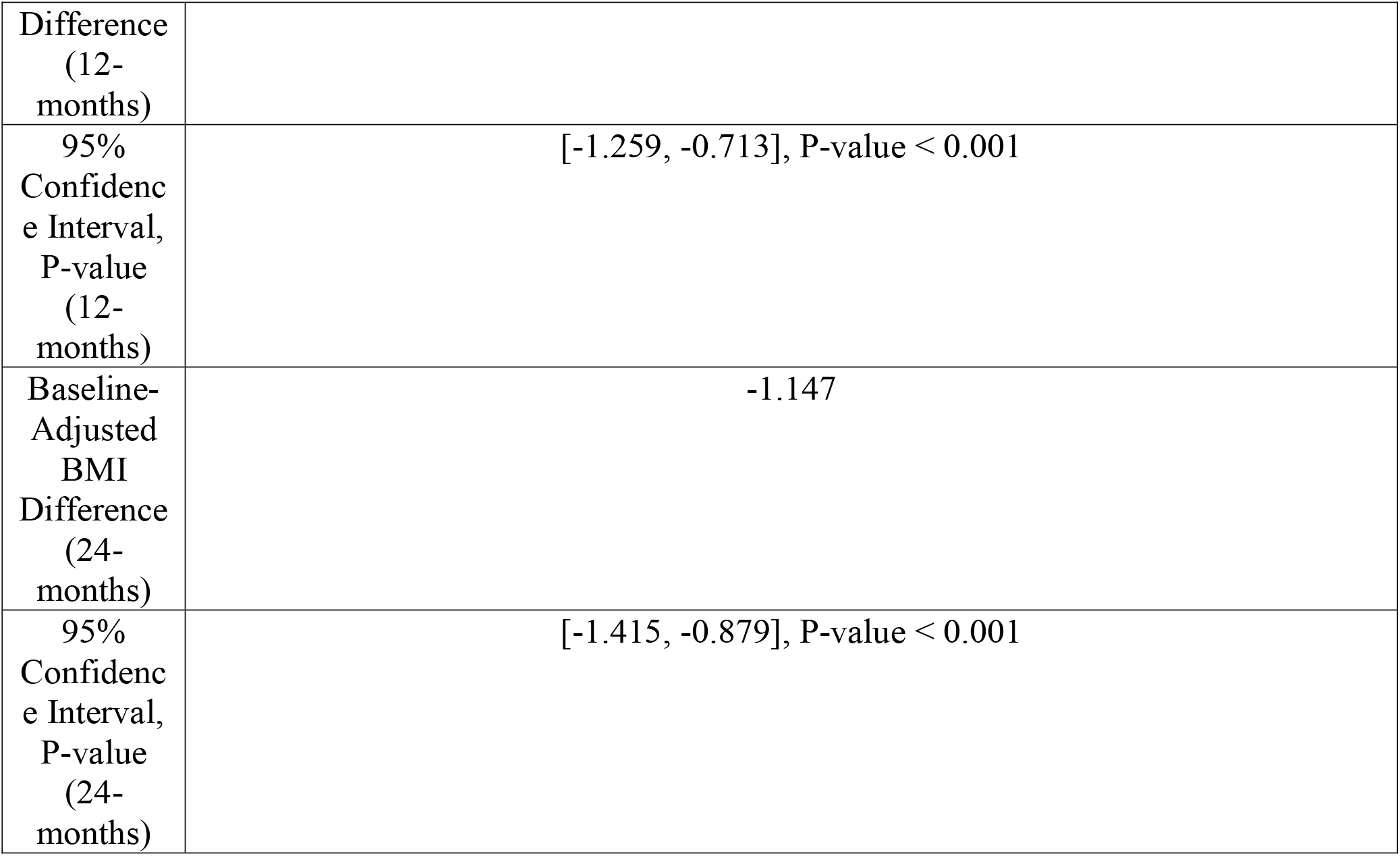
BMI Differences Over Follow-Time Point Comparison Between Tirzepatide Group vs. Control Group.

Follow-up participation varied between groups, with the tirzepatide group showing consistent engagement from 95 individuals at three months to 125 at 24 months, while the control group maintained slightly higher numbers, ranging from 110 to 138 participants. Despite these variations, the absolute BMI differences between groups remained statistically significant throughout the study period, with the tirzepatide group maintaining higher absolute BMI values but showing greater relative reductions over time. This sustained difference, coupled with the progressive baseline-adjusted reductions, suggests a significant effect of tirzepatide on weight management in our IIH cohort compared to standard management alone.

## 4. Discussion

The emergence of GLP-1 receptor agonists has revolutionized the therapeutic landscape for IIH, offering a novel approach that addresses both the metabolic and neurological aspects of the disease [1, 2]. While the efficacy of single-receptor GLP-1 RAs as exenatide has been demonstrated through two clinical studies on IIH patients [8, 10], our study presents the first large-scale investigation of tirzepatide, a dual GIP/GLP-1 receptor agonist [11], as an adjunctive therapy in IIH management. The complex interplay between obesity, metabolic dysfunction, and ICP regulation in IIH suggests that a dual-receptor approach may offer advantages over traditional single-receptor therapies [4, 7].

Our study represents a significant milestone advancement in IIH literature through its utilization of the TriNetX database, enabling a structured propensity score-matched analysis of 435 tirzepatide-exposed patients against 870 controls. This methodological approach addresses several limitations common to previous IIH studies, where sample sizes have typically been restricted. The careful matching process created well-balanced cohorts across demographic and clinical parameters, effectively mitigating selection bias and strengthening the validity of our findings. The rationale for investigating tirzepatide specifically stems from emerging evidence regarding the role of metabolic dysfunction in IIH pathophysiology [4, 12]. Recent research has demonstrated that adipose tissue dysfunction, particularly in the context of obesity, contributes to IIH through multiple mechanisms, including the production of pro-inflammatory adipokines and alterations in hormonal signaling [3, 12].

Our findings demonstrate significant therapeutic benefits across multiple clinical endpoints, with particularly notable improvements in papilledema and visual outcomes. The 67.7% reduction in papilledema risk at three months represents a substantial improvement over outcomes reported in previous GLP-1 RA studies [2, 8, 13]. The sustained improvement in visual outcomes, reaching a 73.9% risk reduction at 24 months, carries significant clinical implications given the potentially devastating consequences of visual deterioration in IIH. The reduction in headache symptoms, while significant, aligns with findings from previous GLP-1 RA studies, suggesting this benefit may be a class effect rather than specific to dual receptor activation [5, 8].

The superior efficacy of tirzepatide observed in our study likely reflects its unique mechanism of action through dual receptor activation. Recent evidence has elucidated the complex role of metabolic dysfunction in IIH, particularly through the action of adipose tissue as an endocrine organ [4, 14-20]. Tirzepatide’s concurrent activation of GIP and GLP-1 receptors may provide more comprehensive regulation of metabolic pathways compared to single-receptor agonists. The mechanisms likely involve modulation of Na+/K+ ATPase activity in the choroid plexus, improvement in hypothalamic function, and reduction in inflammatory mediators [2, 3, 21, 22]. Furthermore, the observed BMI reductions in our study (-1.147 kg/m^2^ at 24 months) suggest that tirzepatide’s weight loss effects contribute to therapeutic benefits through reduced intracranial venous pressure and decreased pro-inflammatory adipokine production [4, 7].

The implementation of tirzepatide in clinical practice requires careful consideration of patient selection and monitoring protocols. Based on our findings and current understanding of IIH pathophysiology, optimal candidates include patients with inadequate response to first-line treatments and those with significant metabolic dysfunction [4, 5, 14, 18]. Regular monitoring should include comprehensive ophthalmological examination, metabolic parameter assessment, and documentation of headache frequency and severity. The integration of tirzepatide into existing treatment algorithms should consider its role as an adjunctive therapy, with particular attention to dose escalation and potential interactions with standard medications such as acetazolamide [2, 8, 13, 23, 24].

Several limitations of our study warrant consideration. The TriNetX database structure precluded detailed analysis of tirzepatide dosing effects and comprehensive medication adherence data. The retrospective design introduces potential for unmeasured confounders, and the lack of standardized ICP measurements across centers may impact the generalizability of our findings. Additionally, variable follow-up periods and incomplete data on concurrent medications represent important considerations in interpreting our results.

Looking forward, our findings establish several critical priorities for future research. Prospective randomized controlled trials comparing tirzepatide with other GLP-1 RAs are essential to confirm our observed benefits [25, 26]. Additionally, investigation of tirzepatide’s direct effects on CSF dynamics and inflammatory markers would enhance our understanding of its therapeutic mechanism. The development of optimized treatment protocols and investigation of combination therapy approaches represent important next steps in maximizing the potential of this promising therapeutic option [25, 26].

## 5. Conclusions

Our findings establish tirzepatide as a promising therapeutic advancement in IIH management, with significant implications for clinical practice and future research directions. The dual GIP/GLP-1 receptor agonism mechanism appears to offer distinct advantages over traditional single-receptor approaches, particularly in addressing the complex interplay between metabolic dysfunction and intracranial pressure regulation. The marked improvements in papilledema and visual outcomes suggest that early intervention with tirzepatide could potentially alter the natural history of IIH, especially in cases refractory to conventional therapies. These results challenge the current paradigm of IIH treatment, suggesting that targeted metabolic interventions may be as crucial as direct ICP-lowering strategies. The sustained therapeutic benefits observed over 24 months indicate potential disease-modifying effects that extend beyond symptomatic relief. However, the variable patient responses and complex interaction with concurrent medications underscore the need for personalized treatment approaches and careful patient selection. Critical questions remain regarding the optimal timing of tirzepatide initiation, dose optimization strategies, and potential synergistic effects with standard therapies. Future evidence should prioritize prospective randomized trials investigating these aspects, alongside mechanistic studies examining tirzepatide’s direct effects on CSF dynamics and neuroinflammatory pathways. The development of biomarkers predicting treatment response and standardized protocols for monitoring therapeutic efficacy would significantly enhance clinical decision-making. As the therapeutic landscape for IIH continues to evolve, our findings suggest that incorporating metabolic modulators like tirzepatide into treatment algorithms could fundamentally transform disease management strategies. This paradigm shift necessitates a reconsideration of current treatment hierarchies and highlights the importance of addressing both the neurological and metabolic aspects of IIH for optimal patient outcomes.

## Data Availability

All used data is available within TriNetX database platform.

## Declarations

## IRB Approval

Waived.

## Ethical Approvals

Waived.

## Consent for Participation

N/A.

## Acknowledgements

Open access funding is provided by the Qatar National Library.

## Data Availability Statement

All used data is available within TriNetX database platform.

